# Association of Nirmatrelvir/Ritonavir treatment and COVID-19 neutralizing antibody titers in a longitudinal healthcare worker cohort

**DOI:** 10.1101/2023.06.19.23291620

**Authors:** Slade Decker, Shaoming Xiao, Carly Dillen, Christina M. Schumacher, Aaron M. Milstone, Matthew Frieman, Amanda K. Debes

## Abstract

Nirmatrelvir/Ritonavir (NMV/r) is used for the treatment of COVID-19 infection. However, rebound COVID-19 infections can occur after taking NMV/r. We examined neutralizing antibodies to the SARS-CoV-2 spike protein before and after infection in people who did and did not take NMV/r to determine if NMV/r impedes the humoral immune response.

## Introduction

In 2021, the Unites States Food and Drug Administration granted Emergency Use Authorization to Nirmatrelvir/Ritonavir (NMV/r) for the treatment of mild-to-moderate COVID-19 infection. Soon after, the United States began providing anti-viral medication at no cost to patients to reduce COVID-19 related hospitalizations and mortality. Reports then emerged of rebound COVID-19 infections, defined as detectable viremia and/or symptoms, two to eight days after completing NMV/r treatment [1-3]. One hypothesis to explain post-treatment rebound infection is that early viral suppression reduces the immune response, including antibody production, thereby predisposing to recurrent viremia and symptoms [4-6]. Prior studies have found that for other viruses, in addition to SARS-CoV-2, early viral suppression can weaken immune response to infection [4-5]. Our objective was to examine neutralizing antibodies to the SARS-CoV-2 spike protein before and after infection in people who did and did not complete an NMV/r treatment course to elucidate whether NMV/r impedes humoral immune response to SARS-CoV-2.

## Methods

### Study Population

Healthcare workers (HW) were consented into an ongoing seroprevalence cohort beginning in June 2020. Approximately every two to six months, HW provided serum samples and completed surveys that captured data including COVID-19 infection dates and NMV/r use. We measured changes in neutralization antibody levels using pre-/post-infection serum samples from each individual meeting inclusion criteria and compared changes in neutralization antibody levels by NMV/r use. HW who received NMV/r (treated) were matched to a HW who did not (untreated). Inclusion criteria included: 1) COVID-19 infection (positive PCR or antigen test); 2) no prior infection or receipt of vaccine dose within 90 days of pre-infection serum collection; 3) had serum collected and stored within 90 days before infection, 4) had serum collected and stored at least 10 days post-infection. Treated HW were matched to untreated HW who had: 1) an infection date +/-14 days of a treated HW; and 2) dates of both the pre- and post-infection serum collection were +/-7 days of the treated HW serum collection. This study was approved by the Johns Hopkins University institutional review board.

### Neutralization assay

Neutralizing antibody titers to the spike protein of SARS-CoV-2 (WA-1) for pre-NMV/r and post-NMV/r (treated) samples were compared with untreated controls to determine if NMV/r had a measurable response on neutralizing antibody response to infection. For the neutralization titer assays (NT), serum samples were heat inactivated at 56°C for 30 min to remove complement and allowed to equilibrate to room temperature prior to processing for neutralization titer. Samples were diluted in duplicate to an initial dilution of 1:10 followed by 1:2 serial dilutions resulting in a 12-dilution series with each well containing 100mL. All dilutions were performed in DMEM (Quality Biological), supplemented with 10% (v/v) fetal bovine serum (heat inactivated, Sigma), 1% (v/v) penicillin/streptomycin (Gemini Bio-products) and 1% (v/v) L-glutamine (2 mM final concentration, Gibco). Dilution plates were then transported into the BSL-3 laboratory and SARS-CoV-2-GFP inoculum was added to each well to result in a multiplicity of infection (MOI) of 0.01 upon transfer to tittering plates. A non-treated, virus-only control and a mock infection control were included on every plate. The sample/virus mixture was then incubated at 37°C (5.0% CO2) for 1 hour before transferring to 96-well flat bottom plates with confluent Vero-TMPRSS2 cells. Titer plates were incubated at 37°C (5.0% CO2) for 24 hours after cells were fixed with 10% neutral buffered formalin (Sigma) for at least 1 hour at 4°C as per BSL3 SOP. Plates were removed from the BSL3 and stained with Hoechest 33342 (Thermo Scientific) diluted 1:2000 in PBS for 10 minutes. Cells were washed with PBS and imaged with a Celigo high content imager (Nexcelom).

### Statistical analyses

The ratio of the half maximal inhibitory concentration (IC50) between matched participants pre- and post-infection were compared using Wilcoxon signed rank test. Statistical significance was defined as p < 0.05. Analyses were performed in R, version 4.2.2 (R Foundation for Statistical Computing, Vienna, Austria).

## Results

Of 1353 HW who reported COVID-19 infection, 65 reported NMV/r use, and 32 met inclusion criteria from which 21 could be matched to a control. We performed neutralization assays on serum from 21 HW treated and 21 untreated with NMV/r. Of the 42 participants, the majority were female (86%) and White (90%). Treated participants were older (median: 50.9; IQR: 44.6-62.0 years) and less frequently female (78% vs. 94%) than untreated participants (median: 35.5; IQR: 32.6-40.9). Treated and untreated participants did not substantially differ by pre-existing conditions or prior exposure to SARS-CoV-2 spike protein.

The pre-infection and post-infection half maximal inhibitory concentrations (IC50) in the treated group were lower than in the untreated group (Figure 1A). The median (IQR) NT of the treated group pre- and post-infection was 980.0 (477.3-2720.3) and 4394.0 (1986.4-7608.4), respectively (Supplemental Table). The median (IQR) NT of the untreated group pre- and post-infection was 1247.5 (433.0, 2032.7) and 7316.0 (4335.0, 10834.0) respectively (Supplemental Table). Of the treated and untreated groups, 67% and 61%, respectively, had an NT greater than or equal to 800 pre-infection, and 100% of both groups had an NT greater than or equal to 800 post-infection. The ratio of IC50 between the treated and untreated groups pre- and post-infection were similar (median: 0.64 and 0.76, respectively, p = 0.7) (Figure 1B).

**Figure 1.**
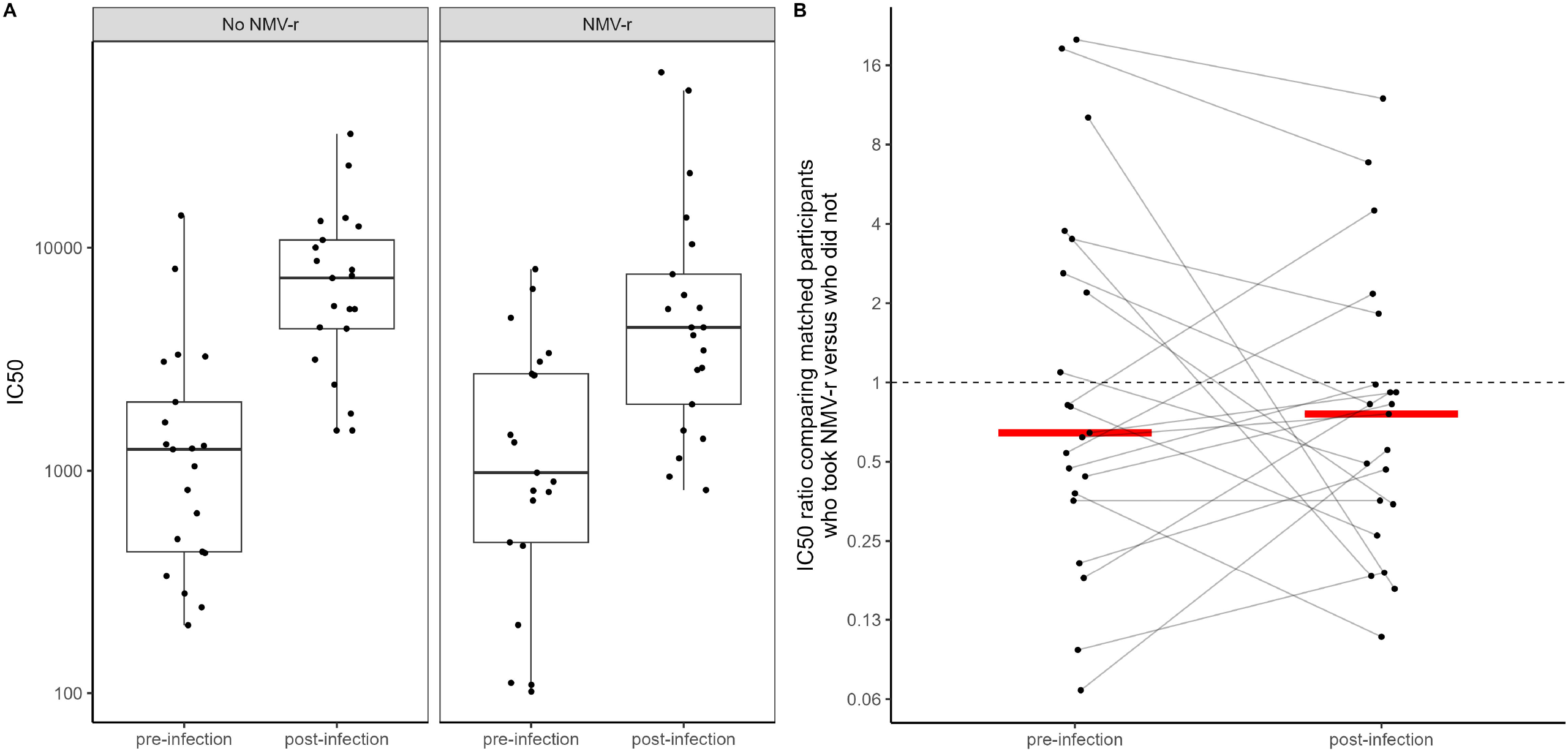
Antibody response to SARS-CoV-2 by NMV/r use among unmatched and matched pairs. Panel A: IC50 pre-infection and post-infection among unmatched participants who took NMV/r (n = 21) and those who did not (n = 21). IC50 values are displayed using a log scale of 10. Panel B: IC50 ratio of matched participants who took NMV/r versus those who did not measured between pre-infection and post-infection serum samples (n = 21). The dashed line represents an equivalent IC50 value (IC50 ratio = 1). IC50 ratio values are displayed using a log scale of 2.

## Discussion

This study demonstrates that taking NMV/r after COVID-19 infection does not impact the host humoral immune response. Our results show that the range between pre-infection and post-infection neutralizing antibody titers for the treated and untreated groups was nearly identical, with median ratios in IC50 values of 0.64 and 0.76, respectively. While this study demonstrates that there is no significant difference in post-infection NT between treated and untreated groups, it highlights that despite high pre-infection NTs, both groups were still infected. Other researchers have attempted to create clarity on the NT threshold of protection, but it remains poorly defined [7].

Our goal was to see if NMV/r has an impact on host humoral immune response during COVID-19 infection and we assessed this by comparing increase of neutralizing antibodies pre-to post-infection. A recent publication accessing immunoglobulin G (IgG) and Omicron-specific neutralizing antibodies in individuals taking NMV/r using only post-infection data reported similar findings, including similar neutralizing antibody levels when taking the medication and when not, with an overall conclusion that NMV/r treatment does not impact adaptive immune response [8]. Our results suggest that concern about early viral suppression reducing immune response to infection should not be a barrier or deterrent to taking NMV/r.

We believe the observed difference in age may be due to indication bias for treatment based on age; older age has been associated with higher risk for severe COVID-19 infection, leading to an increase in older individuals taking NMV/r. The difference in pre-infection and post-infection NT between the treated and untreated groups may be attributed to the age differences across study groups. Prior work has found that compared to older people, younger people produce a higher antibody response to COVID-19 [9-10].

Limitations of this study include a small sample size (n = 42), and a demographically homogenous cohort of HW that limits generalizability, although our sample size is significantly larger than those used in a similar prior study [8]. Secondly, we only performed NT on WA-1 SARS-CoV-2. Despite the antigenic differences between original SARS-CoV-2 and its numerous variants, including Omicron, we tested only WT WA1 strain as that is an identical match to the vaccine strain. Our study population consisted entirely of vaccinated individuals, so using the same strain as the vaccine would result in the most robust responses.

The results of this study show that NMV/r medication does not have a significant impact on humoral immune response post-infection. This finding should reassure those infected with COVID-19 that they can take NMV/r without negatively impacting their humoral immune response. More work is needed to break down barriers that have prevented the acceptance of this drug as an important treatment option for COVID-19. This study examined immune response relatively soon after infection; future research is needed to examine the impact of rebound COVID-19 infections on neutralizing antibody titers, and the rate of anti-SARS-CoV-2 antibody degradation in individuals who take NMV/r compared to those who do not over longer time periods post-infection. Our findings demonstrate that those at high-risk can take NMV/r to reduce the severity and length of mild to moderate COVID-19 infection without detriment to their post-infection humoral immune response.

## Data Availability

All data produced in the present study are available upon reasonable request to the authors.

## Acknowledgements

The authors would like to thank Annie Kuh, LuAnn Rezavi, and other members of the Johns Hopkins Hospital Clinical Immunology Laboratory, Danielle Koontz, Anushree Aneja and Emily Egbert of the Johns Hopkins Division of Pediatric Infectious Diseases, and Matt Courtemanche from the Johns Hopkins University School of Medicine. We would also like to thank Dr. Ralph Baric (UNC Chapel Hill) for providing the SARS-CoV-2-GFP inoculum. Research reported in this publication was supported in part by the National Institute of Allergy and Infectious Diseases of the National Institutes of Health (NIH) under award number K24AI141580 (A.M.), HHSN272201400008C / 0258-0686-4609 (MF and CD), and the generosity of the collective community of donors to the Johns Hopkins University School of Medicine and the Johns Hopkins Health System for Covid-19 research.

