## Supplemental Table 1 for "Association of Nirmatrelvir/Ritonavir treatment and COVID-19 neutralizing antibody titers in a longitudinal healthcare worker cohort"

**Table S1.** Live-virus neutralizing antibody titers (NT) against the vaccine strain (WA-1). Values are in median (IQR).

| Study Group | Pre | Post | Fold Change |
| --- | --- | --- | --- |
| NMV-r | 980.0 (477.3 – 2720.3) | 4394.0 (1986.4, 7608.4) | 3.6 (1.3 – 11.2) |
| No NMV-r | 1247.5 (433.0 – 2032.7) | 7316.0 (4335.0 – 10834.0) | 5.1 (4.0 – 12.8) |
